# Evaluation of non-contact infrared thermometers for COVID-19 fever screening in healthcare workers: A prospective observational study in a tertiary care hospital

**DOI:** 10.1101/2023.12.21.23300100

**Authors:** D. Suresh Kumar, R. Ratheesh, M. Poojadharshini, P. Kalyani, S. Pooja

## Abstract

**Background:** In the fight against COVID-19, efficient fever screening was essential to curb transmission. Fever served as a cardinal symptom, aiding early and timely identification of fever among healthcare workers (HCWs) was crucial. While non-contact infrared thermometers (NCITs) offered non-invasive screening, existing data gaps were present. This study aimed to assess the NCIT effectiveness in HCW fever screening by comparing results with serology and RT-PCR tests, ascertaining their utility in healthcare settings for COVID-19 detection.

**Methods:** This prospective observational study was conducted at a dedicated COVID-19 tertiary care hospital with 250 beds in South India. The study population comprised 736 healthcare workers (HCWs) working in the hospital, and the study was carried out between April 2020 and December 2020. Daily fever screening using non-contact infrared thermometers (NCITs) was performed on all HCWs upon their entry to the hospital. Additionally, serological tests were offered to all HCWs starting from November 2020 to assess prior COVID-19 infection exposure. COVID-19 admissions were closely monitored during the study period to identify hospitalized HCWs with symptoms who subsequently tested positive for COVID-19 using RT-PCR.

**Results:** In this study cohort of 736 HCWs, 44,836 NCIT screenings revealed no fever cases. The serological analysis identified prior COVID-19 exposure in 229 HCWs. McNemar’s test (χ^2^ = 26.27, p < 0.05) emphasized discordance between NCIT and serology. ROC analysis yielded an AUC of 0.500, indicating NCIT’s challenge in distinguishing febrile cases. Additionally, 68 symptomatic HCWs tested COVID-19 positive through RT-PCR, highlighting the role of complementary diagnostics.

**Conclusion:** The failure of NCIT to identify fever cases in our study highlights the importance of incorporating supplementary screening methods and comprehensive strategies in future pandemic preparedness.

## BACKGROUND

In the global battle against the COVID-19 pandemic, effective fever screening has emerged as a pivotal strategy to mitigate viral transmission and prevent potential outbreaks. Fever acknowledged as a cardinal symptom of COVID-19, serves as a crucial indicator of infection, playing a pivotal role in facilitating prompt screening, isolation, and containment—particularly in the absence of vaccination. [1][2][3]

Several studies have underscored the importance of early fever detection among healthcare workers (HCWs), highlighting its potential to mitigate nosocomial transmission and ensure patient safety. HCWs, situated on the frontline of the pandemic, confront an elevated risk of infection due to direct patient interactions. This underscores the urgency of deploying accurate fever screening methods to identify potential carriers of the virus within this critical group. [4][5][6]

Notably, non-contact infrared thermometers (NCITs) offer a non-invasive and efficient means of fever detection, effectively minimizing direct contact and reducing the risk of viral transmission. The versatility of NCITs demonstrated across various settings during different respiratory viral pandemics, extends from bustling airports to healthcare facilities. [7][8][9]

Despite the increasing adoption of NCITs, limited comprehensive data exists, especially from developing nations, regarding their effectiveness for fever detection, particularly among HCWs in varying environmental conditions. [10][11]

## AIM & OBJECTIVE

This study aimed to evaluate the effectiveness of non-contact infrared thermometers (NCITs) for fever screening among healthcare workers (HCWs) in the hospital setting during the COVID-19 pandemic. The objective was to compare the results obtained from NCIT screening with serology and reverse transcription-polymerase chain reaction (RT-PCR) tests to study the utility of NCITs as a fever screening tool in the healthcare setting.

## METHODOLOGY

### Study Design and Setting

This prospective observational study was conducted at a dedicated COVID-19 250-bed tertiary care referral hospital in South India. Convenience sampling was employed to recruit 736 healthcare workers (HCWs) actively working in the hospital between April 2020 and December 2020.

### Fever Screening

Daily fever screening using non-contact infrared thermometers (Model: TOGO TG8818H measuring range 35-42□ (95-107□) manufactured by Lsl tools Pvt Ltd) was conducted on all HCWs upon their entry to the hospital premises. The number of NCIT screening encounters for each HCW was meticulously recorded throughout the study duration, resulting in a total of 44,836 screening encounters.

### Serological Testing

Starting from November 2020, serological tests were offered to all HCWs to assess prior exposure to COVID-19. Only HCWs who voluntarily provided a blood sample were included in the analysis to minimize selection bias. Serological tests were utilized to detect the presence of COVID-19 antibodies, indicating previous infection.

COVID-19 Admissions Monitoring: Throughout the study period, all COVID-19 admissions of HCWs with fever symptoms were closely monitored. Among the HCWs admitted with fever symptoms and tested positive using reverse transcription-polymerase chain reaction (RT-PCR using CoviPath an COVID-19 RT-PCR kit manufactured by Thermo Fisher Scientific), only those individuals were included and analysed to compare the utility of NCITs. (Figure 1)

**Figure 1:**
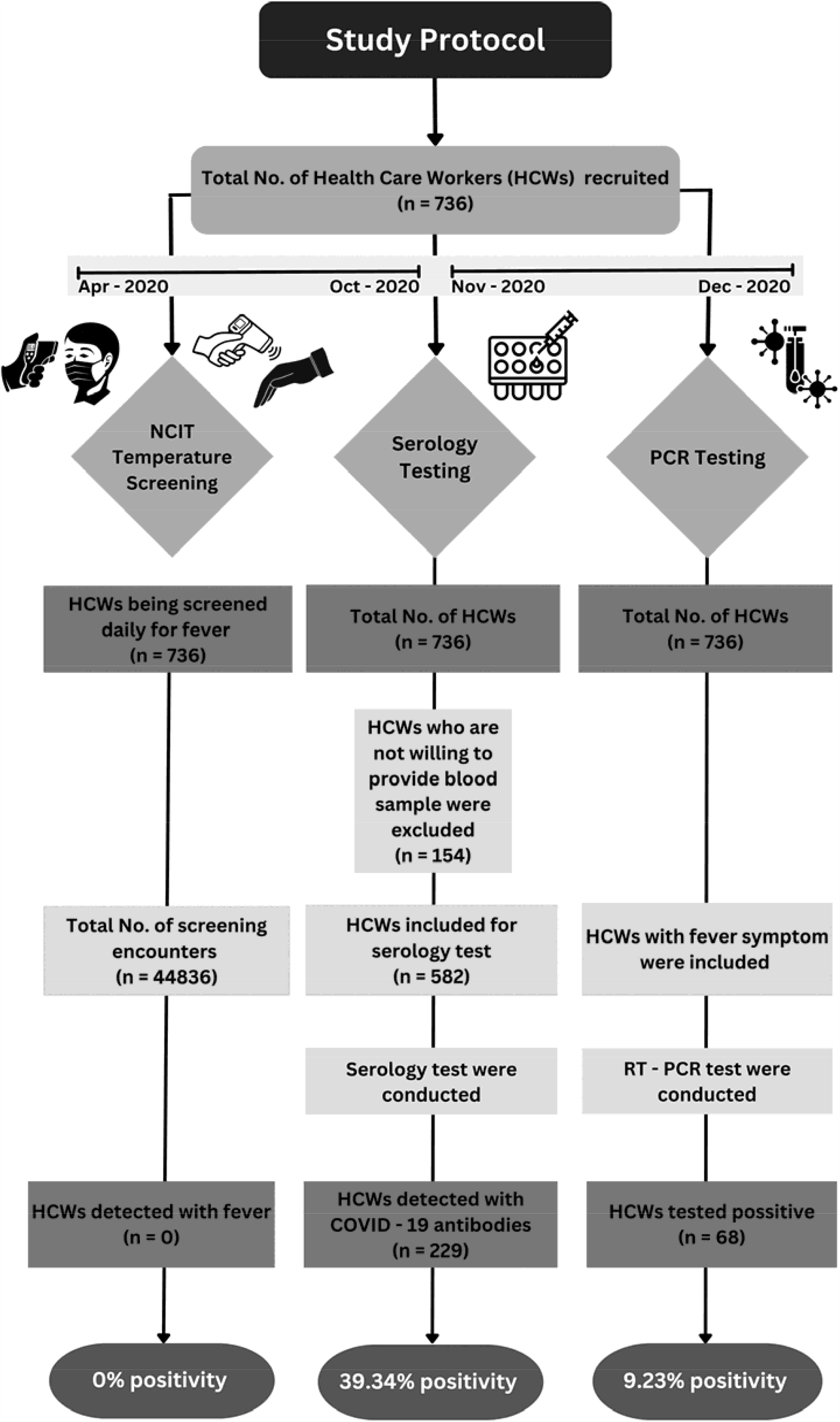
Flowchart interpreting positivity rates of different testing methods in identifying fever among healthcare workers.

## STATISTICAL ANALYSIS

Descriptive statistics were employed to present the number of NCIT screening encounters and serological test results in frequency tables. Concordance between NCIT screening and serology was evaluated using McNemar’s test, generating a χ^2^ statistic and corresponding p-value. Furthermore, the discriminatory capacity of NCITs for fever detection was assessed through Receiver Operating Characteristic (ROC) analysis. The resulting Area Under the Curve (AUC) quantified NCITs’ diagnostic performance. The SPSS software was utilized for all statistical analyses. Significance was established at p < 0.05 for all tests.

Ethical Considerations: Informed consent was secured from all participants, with prior ethical endorsement granted by the institutional ethics committee, confirming the study’s compliance with ethical standards.

## RESULTS

In the cohort of 736 enrolled healthcare workers (HCWs), a comprehensive record of 44,836 non-contact infrared thermometer (NCIT) screening encounters was meticulously documented. Remarkably, no instances of fever detection were recorded using NCITs. In contrast, the outcomes of serological testing revealed 39% (229/582) seropositivity, indicating prior exposure to COVID-19 as given below in Table 1. This disparity in results between NCITs and serology underscores the intriguing complexity of fever screening among healthcare workers.

**Table 1:** Descriptive Statistics of Healthcare Workers and Screening Outcomes.

| Parameter | Total Number | Percentage (%) |
| --- | --- | --- |
| Healthcare workers (HCWs) | 736 | - |
| NCIT Screening Encounters | 44,836 | - |
| Serological Testing | 582 | - |
| Serology Positive cases | 229 | 39.34% |
| RT – PCR Positive cases | 68 | 9.23% |

The concordance between NCIT screening and serology for fever detection was assessed using McNemar’s test, yielding a statistically significant test statistic of approximately 26.27 (χ^2^ = 26.27, p < 0.05). This substantial divergence in the proportions of positive cases identified by these two distinct methods highlights the limited agreement between NCITs and serology in detecting febrile cases among HCWs. These findings underscore the challenges of relying solely on NCITs for fever screening and emphasize the need to carefully consider alternative diagnostic approaches.

The application of Receiver Operating Characteristic (ROC) analysis offered supplementary insights into the discriminative potential of NCITs in fever detection. The calculated area under the ROC curve (AUC) stood at 0.500, signalling a lack of discriminatory capability. This outcome underscores the inherent challenges that NCITs face in accurately differentiating between febrile and non-febrile cases among HCWs within the hospital environment during the COVID-19 pandemic as given below in Figure 2.

**Figure 2:**
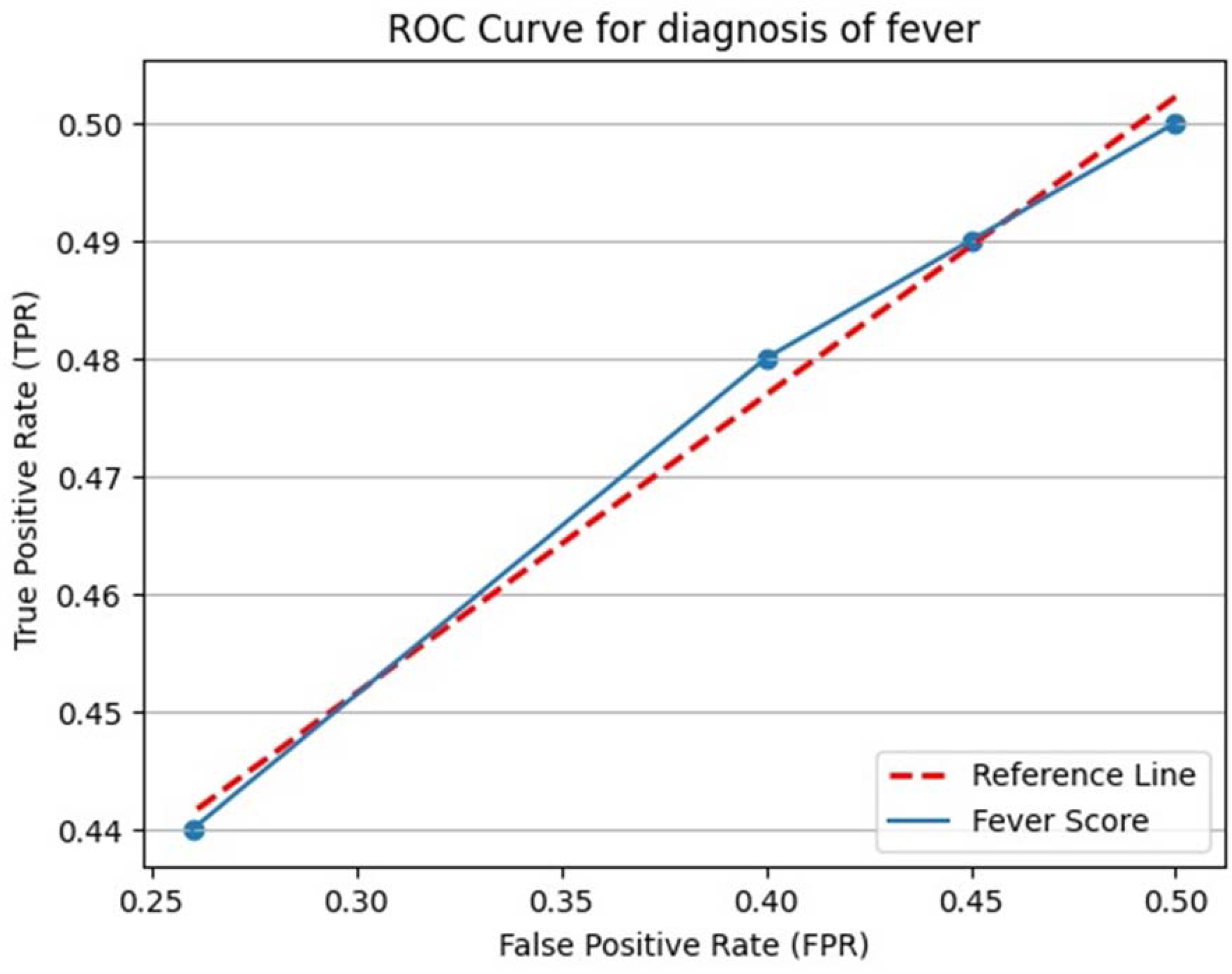
“Fever Detection Evaluation: ROC curve highlighting the discriminative potential of fever detection approaches. McNemar’s test yielded a statistically significant divergence (χ^2^ = 26.27, p < 0.05) in positive case identification between methods. The calculated area under the ROC curve (AUC = 0.50) reflects the challenges of accurately distinguishing febrile cases among healthcare workers in hospital settings during the COVID-19.”

Significantly, the study period witnessed 68 HCWs, displaying fever symptoms, who were subsequently hospitalized and tested positive for COVID-19 through reverse transcription-polymerase chain reaction (RT-PCR) testing. These observations underscore the pivotal role played by complementary diagnostic modalities, such as RT-PCR, in effectively detecting COVID-19 cases, especially among symptomatic HCWs.

## DISCUSSION

We assessed the efficacy of non-contact infrared thermometers (NCITs) for fever screening among healthcare workers (HCWs) during the COVID-19 pandemic. While our extensive NCIT screenings (44,836) failed to detect fever cases among 736 HCWs, serology identified 229 (39%) HCWs with prior COVID-19 exposure, underscoring the disparities between NCITs and serology. McNemar’s test and ROC analysis reinforced these findings, while among HCWs with fever symptoms, 68 tested positive for COVID-19 via RT-PCR. Our study emphasizes the need for supplementary screening methods and comprehensive strategies in pandemic preparedness, alongside recognizing the limitations of NCITs as standalone fever screening tools for COVID-19.

Our study’s findings of no fever cases detected through extensive NCIT screenings align with recent investigations of Nsawotebba’s study among truck drivers with a sensitivity of (1.2%) for temperatures exceeding 37.5°C, with a low 9.9% sensitivity of thermal screening and to Khan S et al. finding of only 16.13% sensitivity and an AUROC of 0.67 at temperatures ≥37.5°C. These congruent outcomes strengthen the conclusion that standalone thermal screening is inadequate for robust COVID-19 screening. [12][13]

In the context of serological testing, Srihita M et al.’s (2021) study among healthcare workers revealed 42% seropositivity within a similar timeframe. Our serological testing yielded a comparable 39% seropositivity, further underscoring significant prior COVID-19 exposure. The consistency between our findings and these studies supports the broader understanding of the limitations and implications of using non-contact infrared thermometers for COVID-19 screening in healthcare settings. [14]

However, our study diverged notably from Hussain et al.’s study, revealing substantial disparities in the sensitivity of non-contact infrared thermometers (NCITs) for fever detection among healthcare workers, attributed to variations in screening distance and temperature thresholds. While Hussain et al. achieved 70.9% sensitivity through NCIT adjustments, our study employed distinct parameters, resulting in lower sensitivity. These differences underscore the importance of considering multiple factors when assessing NCIT efficacy for fever screening in diverse healthcare settings. [15]

In contrast, Fan Lai and colleagues emphasized the inadequacy of single NCIT measurements for fever screening, advocating triple neck measurements as the standard, and highlighted NCIT accuracy decline below 18°C due to environmental conditions. This comprehensive approach, addressing temperature variations, enhances our comprehension of NCIT intricacies and limitations. [16]

Additionally, our investigation demonstrated substantially higher seropositivity (39%) among healthcare workers compared to Ranjana Hawaldar et al.’s study reporting a 7.82% seroprevalence in Central India. These divergent findings may result from disparities in study timelines and serological methodologies. [17]

This study offers valuable insights into the constraints of relying solely on non-contact infrared thermometers (NCITs) for COVID-19 fever screening among healthcare workers. It underscores the significance of employing complementary screening methods. However, the study’s limitation lies in its single-site focus, potentially limiting the applicability of findings to other healthcare settings. Moreover, the investigation did not extensively explore NCIT accuracy across different ambient temperatures and distances, which could have further enriched the study’s outcomes.

## Conclusion

In conclusion, our study highlights the limitations of standalone non-contact infrared thermometers (NCITs) for COVID-19 fever screening among healthcare workers, underscoring the importance of supplementary screening methods. The disparity between NCIT-based detection and serological findings urges a comprehensive approach in future infection pandemics. As healthcare facilities prepare for future pandemics, our insights prompt a reconsideration of screening protocols that acknowledge the strengths and limitations of different screening tools.

## Data Availability

All data produced in the present work are contained in the manuscript

## Ethical statement

Ethics approval was obtained from institutional ethics committee of Apollo hospitals, Chennai, Tamil Nadu, India (IEC-BMR App No: AVH-C-S-009/07-23).

## Informed consent

The participant has consented to the submission of the article to the journal.

### Funding

The authors received no financial support for the research, authorship, and/or publication of this article.

---

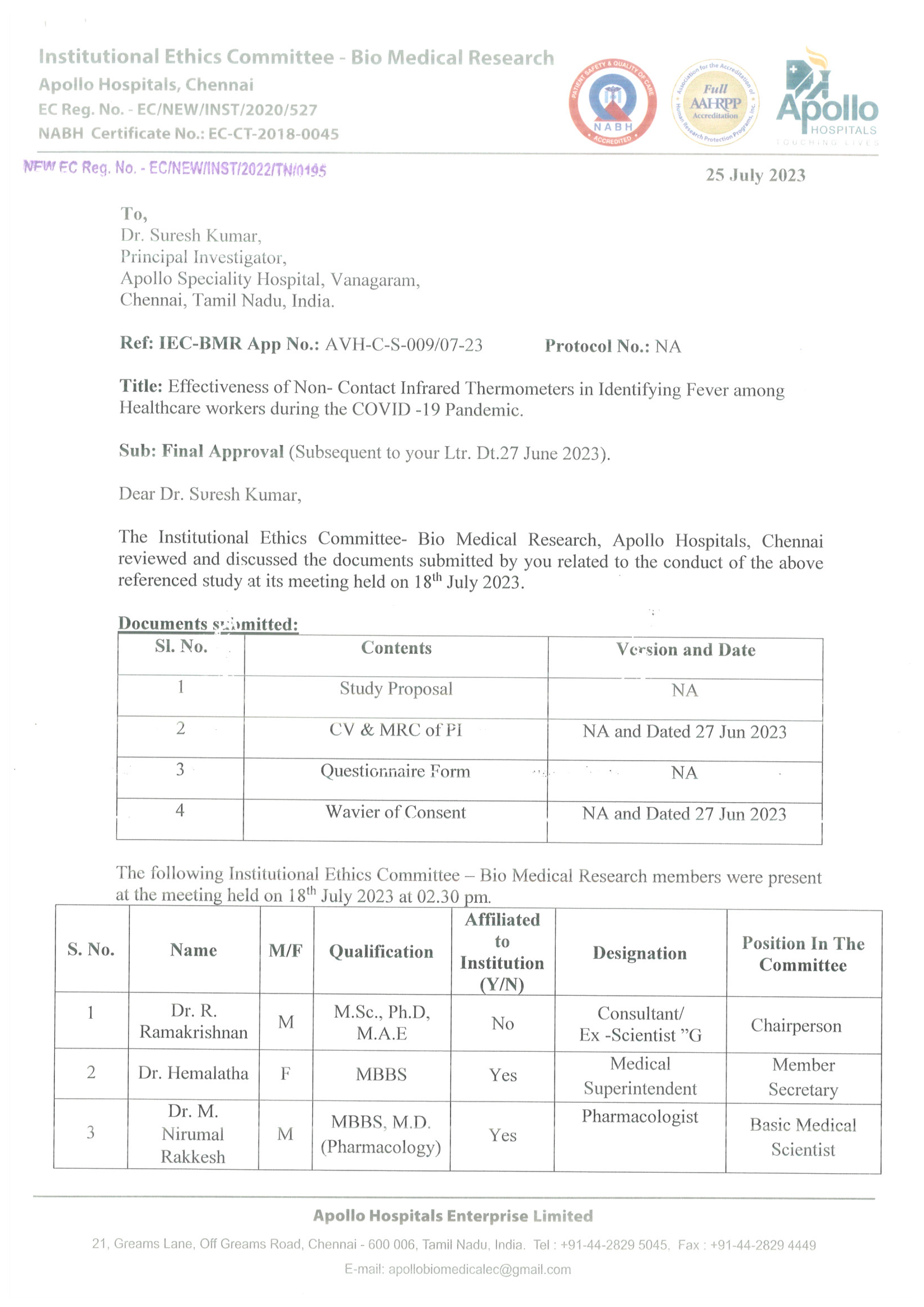

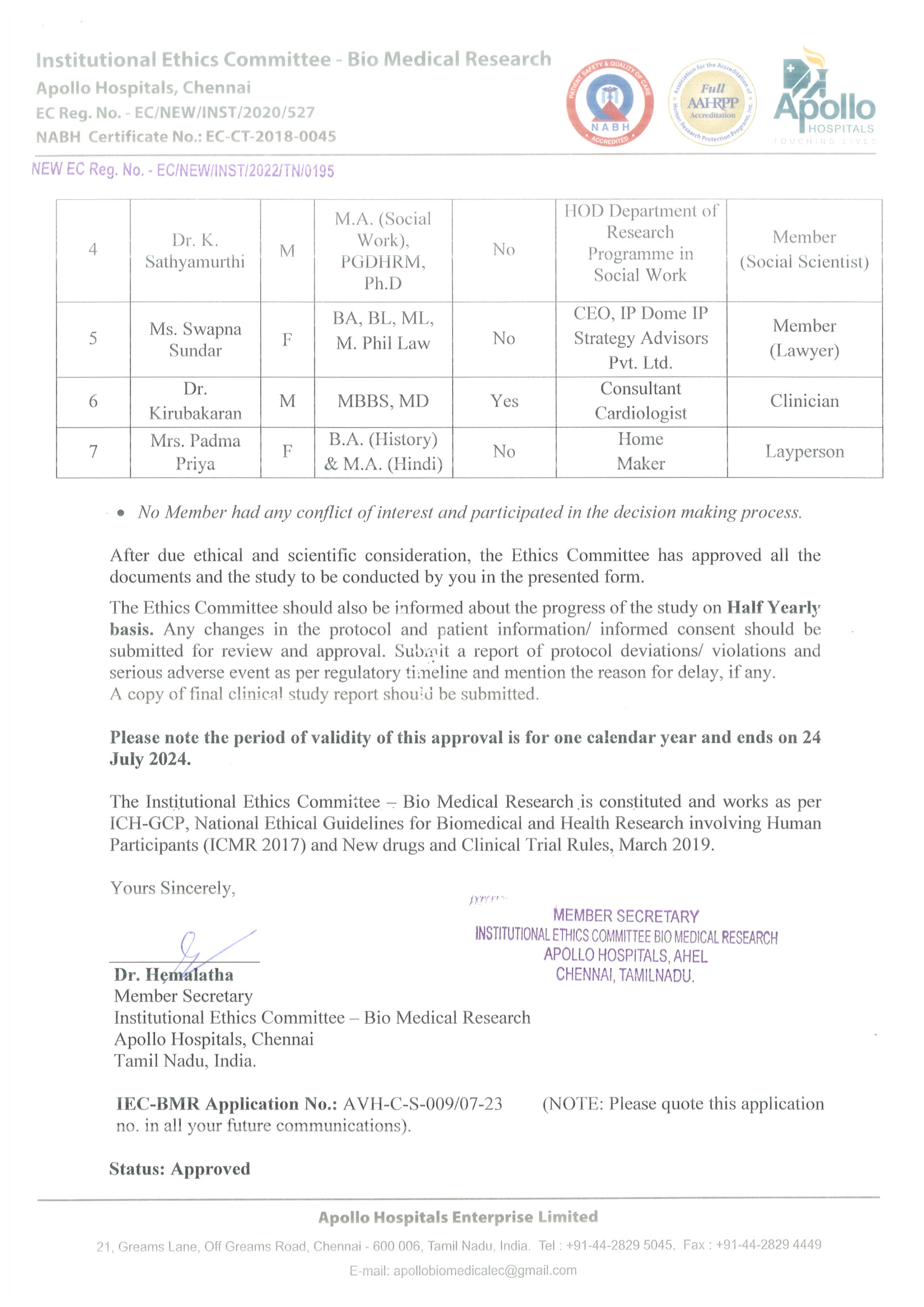

